# Developing best practice rare disease diagnostic care models in a real-world rural/regional setting

**DOI:** 10.1101/2025.07.24.25332141

**Authors:** Lauren S. Akesson, Sanjoti Parekh, Andrew Alderdice, Hannah Jackson, Lynda Bain, Alice Dudgeon, Linda J. Williamson, Brianna L. Akesson, Genevieve Say, Miranda J. Kellett, Rachel Pope-Couston, Mathew J. Wallis

## Abstract

**Background:** Although individually rare, collectively rare diseases are more common than diabetes mellitus. The diagnostic odyssey for rare diseases is typically prolonged.

**Aim:** We aimed to hear the voice of rare disease stakeholders to provide evidence for strategies to improve rare disease care in Tasmania, Australia.

**Methods:** Mixed methods research with surveys, focus groups, and interviews captured the voice of healthcare consumers with rare disease, their families, and their caregivers, as well as consumer advocates, community reference group, rare disease researchers, and healthcare professionals.

**Results:** We undertook 1,014 stakeholder consultations. Participants described elements of the diagnostic pathway that worked well, as well as barriers to diagnosis. Those affected by rare disease and their caregivers emphasised the need for healthcare providers to listen and respond to consumer concerns. Collectively, stakeholders reported that challenges in rare disease diagnosis in Tasmania include difficulties accessing timely primary and specialist health care, low awareness of rare disease among health professionals in Tasmania (including how to identify people with possible rare disease and what to do next), difficulty in accessing diagnostic testing, cost and geographic barriers in accessing diagnostic care, a lack of epidemiologic information on rare disease in Tasmania and the breakdown in care and support from the health system when transitioning from paediatric to adult services.

**Discussion:** Many participant concerns that fall under the system responsibility of the Tasmanian Government could be addressed by establishing a Tasmanian Rare Care Centre. The centre could adopt principles from cancer care, such as patient-centred care, safe and quality care, multidisciplinary care, supportive care, care coordination, communication, and research and clinical trials. Other concerns, such as the equitable implementation of emerging diagnostic technologies and the education and training of non-genetics practitioners, will require national leadership and interjurisdictional cooperation to address.

SUMMARY BOX
**The Known**
Rare diseases are a global health priority. The diagnostic odyssey is often prolonged, affecting rare disease care.
**The New**
Stakeholder consultation revealed that individuals affected by rare diseases and their families and caregivers often feel that they are not listened to, that their care is fragmented, and that non-genetics healthcare providers have limited awareness and understanding of rare disease.
**The Implications**
Many concerns raised by rare disease stakeholders could be addressed by establishing a Tasmanian Rare Care Centre.

## INTRODUCTION

Rare diseases are a global public health priority,^1^ affecting a significant number of people despite their individual rarity.^2^ In Australia, rare diseases are defined as conditions with fewer than five cases per 10,000 people; in total, they impact an estimated 40,000 Tasmanians.^3,4^ Families in rural and regional areas face greater challenges in obtaining timely diagnoses and accessing care, often enduring years-long diagnostic odysseys.^5^ The Australian Government’s National Strategic Action Plan for Rare Diseases, released in 2020, aims to improve healthcare equity, particularly in underserved areas like Tasmania, where all residents are classified as living in regional, rural, or remote areas (Australian Statistical Geographic Standard, Volume 6, 2021).

Addressing these challenges requires improvements to diagnostic pathways, healthcare access, genomic medicine implementation, and community involvement in genomic research.^6^ Primary care remains the backbone of Tasmanian health systems,^1^ yet geographical isolation and practitioner availability limits healthcare options.^7^ The rare disease diagnostic odyssey is often prolonged, measured at approximately 5 years.^8,9^ Misdiagnoses and delays in care are common,^10^ but advances in rapid genomic sequencing offer hope, especially for critically ill children^11-13^ and adults,^14^ including during pregnancy.^15^ Nevertheless, challenges in rare disease diagnosis limit equity for families accessing healthcare: existing genetic healthcare pathways favour urban areas,^16^ increasing inequity for rural families. Recognising this, the Tasmanian Clinical Genetics Service, supported by Australian Genomics, sought to understand rare disease diagnostic pathways in regional Australia and develop recommendations and new models of care to shorten. The diagnostic odyssey and improve rare disease care. This study employed a mixed-methods approach to identify problems and develop solutions, addressing a methodological gap in the literature.

## METHODS

### ETHICAL CONSIDERATIONS

This study was approved by the University of Tasmania Human Research Ethics Committee (H002780). With the primary aim of this study being to collect the viewpoints of rare disease stakeholders, data gathering and analysis was designed to be independent of the public health service, to give participants added confidence that their responses would not adversely affect their healthcare. To do this, the Tasmanian Clinical Genetics Service engaged the independent firm Abt Global. Surveys were conducted by Abt Global and focus groups and interviews were conducted at neutral sites and not at health facilities. Raw data was analysed by the consultants at Abt Global with aggregated data provided to the Tasmanian Clinical Genetics Service to preserve participants’ anonymity.

### OVERALL STUDY DESIGN

This project employed mixed methods research with thematic analysis, conducted by Abt Global and overseen by the Tasmanian Clinical Genetics Service as part of the Tasmanian Rare Disease Diagnostic Pathways Project, gathering insights from key stakeholders, including individuals, families, and caregivers affected by rare disease, consumer advocates, health professionals, and researchers. The project aimed to understand how different stakeholders experience rare disease diagnostic pathways in Tasmania and engage those stakeholders in identifying and developing practical system-level solutions to overcome diagnostic barriers.

We used qualitative surveys, focus groups, and semi-structured interviews to capture diverse stakeholder perspectives. We developed 2 standardised surveys: for individuals affected by rare disease and their caregivers, and another for health professionals, researchers, and consumer advocacy groups. Focus groups and interviews with individuals and caregivers followed one interview guide, while a separate guide was used for health professionals, researchers and consumer advocates. These sessions were conducted by representatives from Abt Global who ensured flexible timing and locations to support participant access to the sessions. Abt Global anonymised all responses and returned aggregated results and draft data analysis and interpretation to the Tasmanian Clinical Genetics Service research team for review. Abt Global and the Tasmanian Clinical Genetics Service then worked collaboratively with members of a Community Reference Group and the Tasmanian Rare and Undiagnosed Disease Network of genomic clinicians and researchers to develop possible system responses to improve rare disease diagnostic care in Tasmania.

### PARTICIPANTS

#### 1 Key expert interviews with rare disease and broader health system leaders

Six key informant interviews were conducted prior to the community engagement, with the purpose of: (i) ensuring the evaluation team had a sound contextual understanding of the rare disease sector in Tasmania and Australia, and how it is situated within the Tasmanian health system; (ii) initial identification of the key barriers to rare disease diagnosis in Tasmania and Australia more broadly; and (iii) informing the key questions for the community consultation processes (surveys and direct consults) and/or feasible approaches to the engagement of stakeholders and maximising participation.

#### 2 Community Reference Group interviews

The Community Reference Group established by the Tasmanian Clinical Genetics Service is a group of six individuals who have lived experience of a rare disease (either as an affected person or as a relative or caregiver). Five of these members participated in one-on-one interviews and provided insights into the diagnostic journey.

#### 3 Patient/Carer engagement

Individuals across Tasmania affected by rare disease and their caregivers were invited to participate in an online survey. The survey further invited these individuals to participate in a follow up focus group or individual interview if desired.

##### Survey

We developed an online survey, the link to which was circulated via the Tasmanian Department of Health’s Facebook page and through rare disease advocacy organisations. This survey was the first step in obtaining information from patients with rare disease and their relatives or caregivers. There were 880 valid responses to the survey (significantly more than the expected 150 responses).Responses were anonymous unless individuals chose to provide contact details via the survey form to receive an invitation to participate in follow up focus groups or interviews.

##### Interviews

Participants who provided their contact details for follow up consultations (n=248/28%) were emailed an invitation with 16 online or face-to-face (Devonport, Launceston or Hobart) focus group options (morning, afternoon and evening over 5 days). The option of a 1:1 interview was also offered to those who could not attend and/or did not want to participate in a focus group. From 248 invitations, 63 people participated in a focus group and 8 in one-to-one interviews. An additional 3 people participated having become aware of the consultations through an advocacy organisation. A total of 74 individuals participated in the follow up consultations.

The areas of focus for the consultations were: i) Experience and journey of the health condition; diagnosed or undiagnosed; ii) Barriers and enablers in the diagnostic journey; iii) Cost associated with getting the diagnosis and other cost related barriers; and iv) Interactions with the health system and quality of care.

#### 4 Clinician/Researcher engagement

Clinicians and researchers were engaged through a survey and/or interviews.

##### Survey

We conducted an online survey for health professionals and genomic researchers in Tasmania. The survey was disseminated via the Tasmanian Department of Health’s Facebook page and health professional bodies, such as Primary Health Tasmania’s newsletter. There were 31 responses to this survey, and the profession of these participants was not identified in reported data to the Tasmanian Clinical Genetics Service to preserve anonymity.

##### Interviews

Clinicians and researchers who play a critical role in the rare disease field were nominated by key stakeholders; 15 participants provided their input from various aspects including; a professional within the system (enablers and barriers), their observation and experience of the patient’s perspective of the diagnostic journey, and the quality of life for a patient during and beyond diagnosis. This group of stakeholders provided input into i) System enablers and barriers for clinicians/researcher in supporting early diagnosis; ii) Diagnostic care pathways that patients follow; iii) Enablers on the diagnostic pathway; iv) Barriers, gaps, issues on the diagnostic pathway; and v) Prioritising and addressing the barriers/issues of the diagnostic pathway.

### 5 Advocacy Organisations Interviews

From 15 rare disease advocacy organisations located in Tasmania that were invited to participate, 8 organisations provided insights into the patient journey and a patient perspective on receiving care. The questions focused on areas such as understanding the patient journeys, barrier and enablers in patient journeys, cost of getting a diagnosis and treatment, and supports required during the diagnostic journey.

## DATA ANALYSIS

Surveys were conducted electronically using Qualtrics software (Qualtrics, Provo, UT, USA). Focus groups and interviews were recorded through manual notes (no audio recordings were made to protect the anonymity of participants). Manual notes were digitally transcribed by 2 investigators (SP and AA) and checked by a further 6 research assistants. Data was thematically analysed^17,18^ by 2 investigators (SP and AA) and presented in aggregate form to preserve anonymity of respondents.

## RESULTS

### WHEN THE DIAGNOSIS GOES WELL

Stakeholder responses highlighted certain factors that contributed to a positive diagnostic experience. These included personal attributes such as health literacy and the strong commitment of families to pursuing a diagnosis. Emotional and mental health support, from family, friends, or others with rare diseases, was highly valued, as was practical assistance like transport. Coordinated support from healthcare providers helped individuals and caregivers navigate the health system and access services. Relationships with healthcare providers were a key factor, including strong trust with a consistent primary care provider, and a primary care provider who listens and is also committed to finding a diagnosis. Healthcare providers with a multidisciplinary team approach were perceived as helping the diagnosis go well.

### WHEN THE DIAGNOSIS IS CHALLENGING

Rare disease stakeholders identified several factors that contributed to a challenging diagnostic journey, many relating to health practitioner and system-level factors. A common concern was limited knowledge of rare diseases among primary care providers. Stakeholders acknowledged and agreed that comprehensive knowledge of rare conditions is unrealistic to expect from general practitioners. Stakeholders did expect, however, that primary care providers should be able to identify when a person may have a rare disease and know what to do next. More critically, individuals with rare disease and their caregivers described poor listening skills, disbelief in patient-reported symptoms, and a lack of empathy from some clinicians in all specialties.

Systemic challenges included primary care provider shortages, particularly in rural areas, limited access to local specialists and diagnostic tools, long wait times to see a specialist (specialties commonly identified were paediatrics, neurology, and rheumatology), and poor communication and coordination between healthcare providers. Fragmentated diagnostic pathways disrupted continuity of care. Transition from child-specific to adult services was seen as particularly challenging.

Travel, especially interstate travel, was frequently raised with impacts on personal finances, work, family, and friends, and the need for some families to relocate interstate to access specialist expertise. Financial barriers to diagnosis, including out of pocket expenses, was thought to increase inequity. Key challenges to rare disease diagnosis are summarised below (Figure 1).

**Figure 1.**
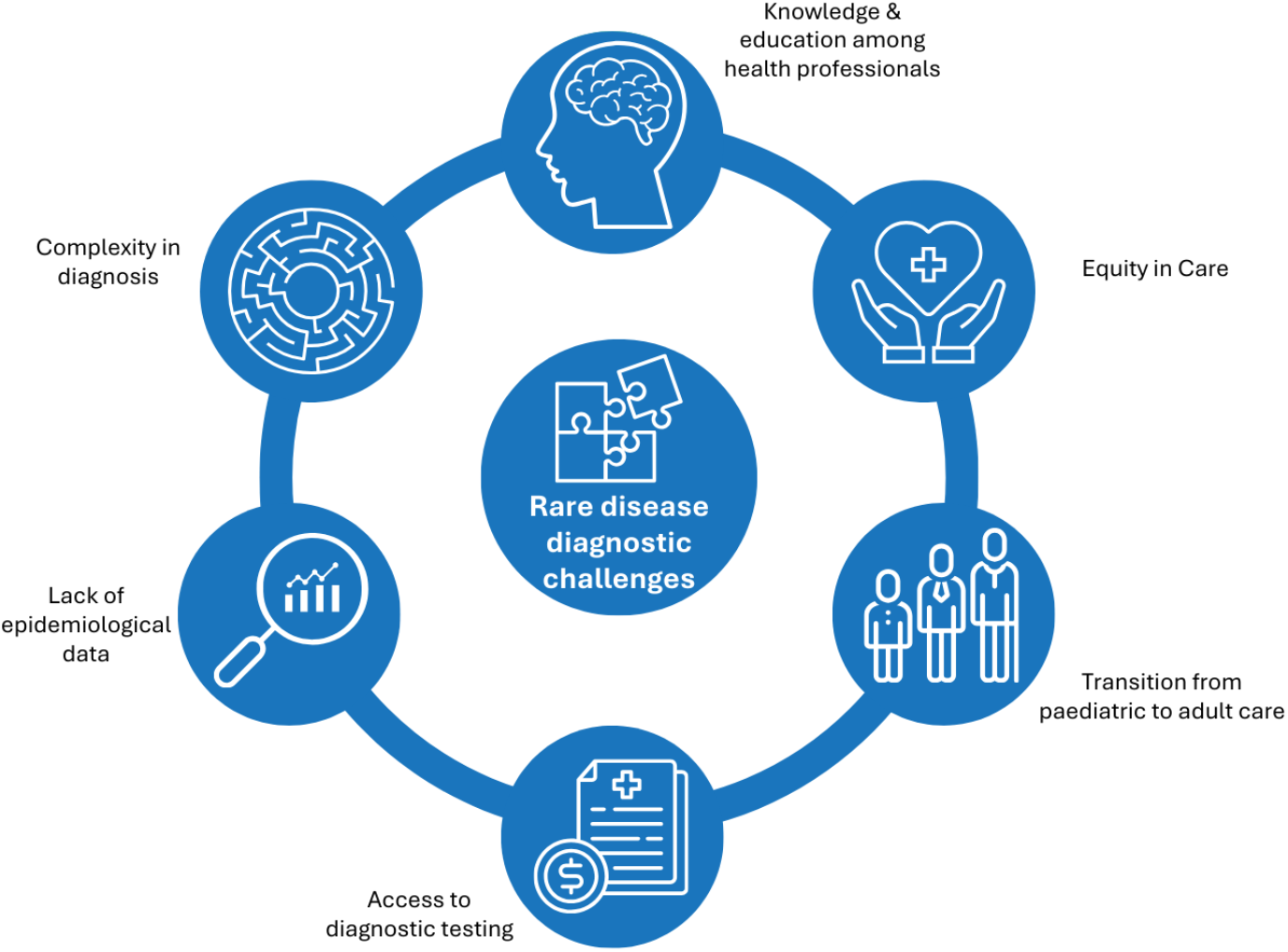
Key challenges in rare disease diagnostics

### WHAT DO INDIVIDUALS AND CAREGIVERS SAY THEY NEED?

The needs of individuals and caregivers affected by rare disease can be categorised into three areas: healthcare provider factors, access to information and education, and support systems. In terms of diagnostic testing, the primary concern expressed was the desire for a shorter timeframe between testing and diagnosis.

Individuals with rare disease and their caregivers emphasised the importance of healthcare professionals who actively listen, consider all symptoms, and include the broader context such as family history. In particular, they valued practitioners who are committed to researching the condition both before and after the diagnosis. They value practitioners who are willing to pursue further investigation, refer for specialist assessment or testing as required and maintain clear communication with families. Effective collaboration, communication and coordination among healthcare providers was also highlighted as crucial.

There was a strong demand for accessible, reliable information about rare diseases, as well as improved access to support. This includes connections to peer networks, access to health, social and disability services (including the National Disability Insurance Scheme), advocacy support and referrals to local support networks within Tasmania.

### SUMMARY: BARRIERS TO RARE DISEASE CARE INCLUDING DIAGNOSIS

Barriers to accessing rare disease care and diagnosis in Tasmania include both challenges that are unique to the state, and those shared with rare disease communities globally (Table 1). Tasmania’s population is significantly dispersed, with two-thirds of the population residing outside Greater Hobart (the Tasmanian capital city, population 247,086).^19^ Greater Hobart and Greater Launceston are considered ‘inner regional’ by the Australian Statistical Geography Standard (ASGS Remoteness Structure), with the remaining Tasmanian regions considered ‘outer regional’, ‘rural’, or ‘remote’. This geographic remoteness was frequently cited by stakeholders as a significant barrier to rare disease care as it contributes to limited access to healthcare professionals.

**Table 1.**
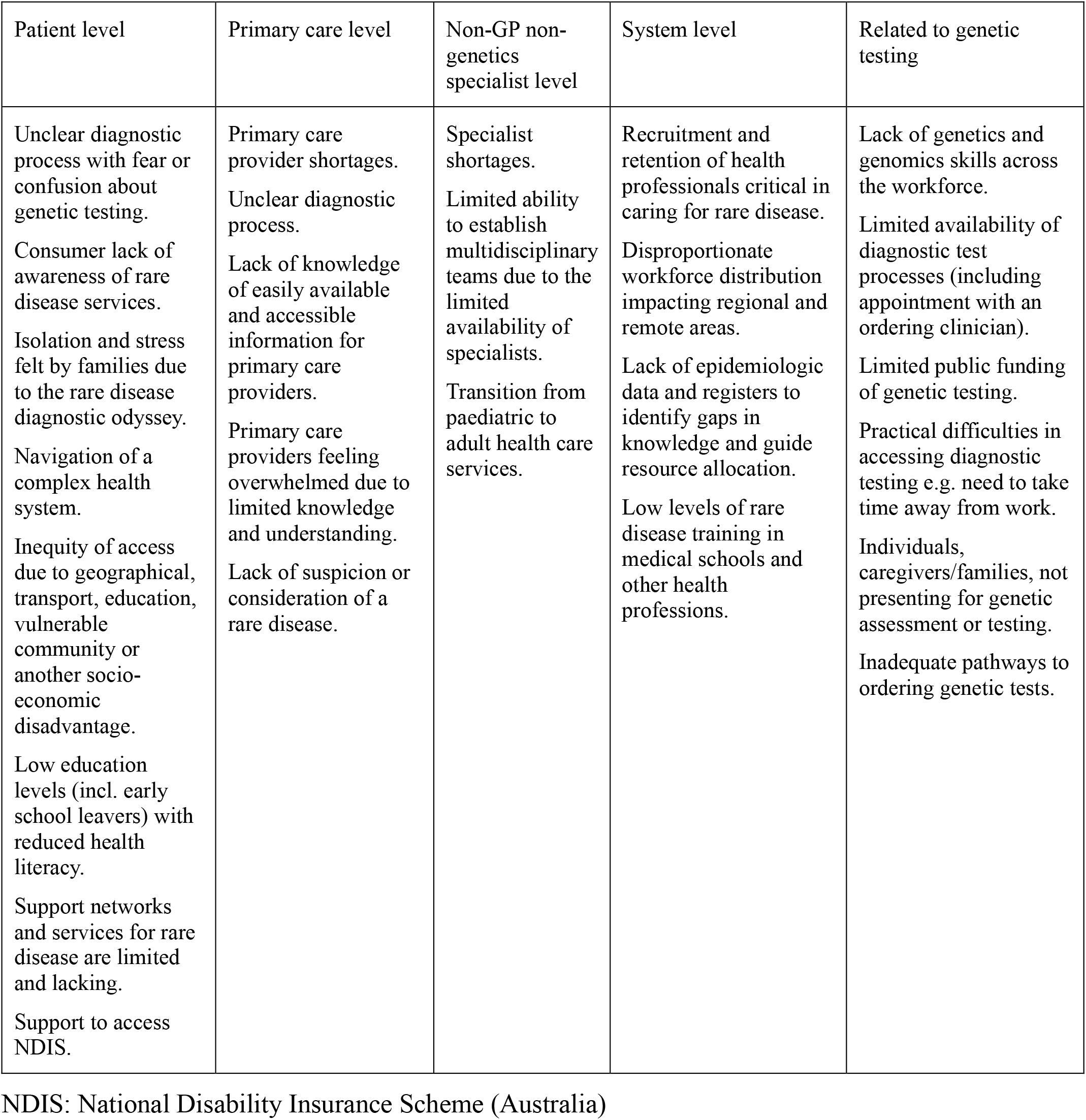
Barriers to rare disease diagnosis.

Individuals living with rare diseases, along with their families and caregivers, expressed that receiving a rare disease diagnosis can feel similar to receiving a terminal diagnosis. They recommended that such diagnoses be delivered with the same level of care and sensitivity, ideally in a family meeting (with consent), supported by access to a counsellor or social worker. Families should also be provided with clear information about the condition, available symptom management options, and guidance on how the diagnosis may impact the mental health of the individual, their family, and caregivers. In essence, they called for a holistic, compassionate approach to diagnosis and care.

## DISCUSSION

The National Strategic Action Plan for Rare Diseases prioritises the development of models of care through ‘establish[ing] standards for care and support that are integrated and incorporate clear pathways throughout all systems’.^20^ Ideally, ‘these [should be] informed by clinical and consumer rare disease experts and that such consultation informs policy development’.^20^ Our mixed methods research builds on the foundations of this consumer consultation by addressing the contextual nuances of Tasmanian healthcare.

The findings from this study led to a series of recommendations based on stakeholder consultation, a literature review of best practice rare disease diagnosis and management, and a review of the current diagnostic pathways in Tasmania. Draft recommendations were developed by Abt Associates and the Tasmanian Clinical Genetics Service and reviewed by the Tasmanian Clinical Genetics Service Community Reference Group, and the Tasmanian Rare and Undiagnosed Disease Network members.

Central to most recommendations for action at the state level was a proposal to establish a **Rare Care Centre** (Table 2). A typical model includes multidisciplinary clinical teams providing coordinated diagnostic and post-diagnostic care, incorporating clinical geneticists and genetic counsellors, specialists, primary care clinicians, nurses, and allied health professionals. These centres provide a dedicated service for the management of complex cases that require coordinated care from multiple specialties. Such a service commonly facilitates access to targeted treatments including clinical trials and can facilitate and coordinate patient and family/carer access to non-clinical support services. Such non-clinical services and supports can be essential for patients and families facing the significant challenges of living with complex rare disease. Underpinning principles of care pathways for such a centre include patient-centred care, safe and quality care, multidisciplinary care, supportive care, care coordination, communication, and research and clinical trials (Figure 2). Such a centre could conduct outreach and raise awareness of rare disease, and the availability of the Tasmanian Clinical Genetics Service among the public and health professionals in Tasmania; and could provide non-genetics health professionals, and individuals with rare disease and their families/caregivers with information and support in relation to both rare disease diagnosis and ongoing care.

**Table 2.** Recommendations for Best Practice Diagnostic Care for Rare Disease.

|  | What does it look like? | How can we achieve this in Tasmania? | What needs to happen on a national level? |
| --- | --- | --- | --- |
| Accessing information | Information is readily available on the underlying causes, mechanisms and clinical presentations of rare diseases for both health professionals and patients and their families and carers. | Establish a Tasmanian Rare Disease support group.<br>Promote existing information resources to both patients and clinicians.<br>Establish a Tasmanian Rare Care Centre. | Establish a national platform or program to encourage information sharing. |
| Accessing the health system | Patients have access to timely primary and specialist healthcare when needed and are supported in navigating the system. | Recruit and retain more clinicians.<br>Improve access to clinicians in rural and regional areas via telehealth.<br>Establish care-coordination for patients with rare diseases, including help accessing associated supports.<br>Facilitate access to rare disease specialists in other jurisdictions. | Increase number of training places for GPs and specialists across Australia. |
| Knowledge and attitudes of clinicians | Health professionals are well informed about rare diseases and | Conduct training and awareness raising to GPs on rare disease and public clinical genetics services. | Improve education on rare diseases and genetics provided to |
|  | available diagnostic and referral pathways.<br>Health professionals listen to their patients and take proactive steps to seek a diagnosis. | Develop and promote resources to guide clinicians in identifying and seeking diagnosis of rare diseases.<br>Develop rare disease referral pathways for inclusion in GP Health Pathways portal. | medical undergraduates and in specialist training. |
| Coordination and integration of care | Patients have access to holistic, coordinated, multidisciplinary care. | Establish a Tasmanian Rare Disease Multi-Disciplinary Diagnostic Clinic.<br>Establish a Tasmanian Rare Care Centre.<br>Promote use of electronic records systems. | Work with primary care software vendors to improve rare disease patient identification and management. |
| Timely and appropriate investigations | Patients have access to timely and affordable diagnostic testing and investigations recommended by appropriate clinical advice and available to all relevant treating clinicians. | Improve local workforce and laboratory capacity to undertake genetic and genomic analysis.<br>Ensure electronic records systems are used to safely document and share patient records. | Increase the range of genetic and other relevant diagnostic tests supported through the national public health system. |
| Accessing Research and Clinical Trials | Families can access research relevant to their condition, supported by local clinicians. | Establish a Tasmanian Rare Disease Registry.<br>Provide ongoing support for translational research participation in relation to rare diseases in the Tasmanian Health Service. | National funding for translational genomic research should specifically support the inclusion and participation of patients, researchers, and clinicians in rural and regional Australia. |
| Accessing Support | Patients and their families and carers can access emotional and mental health support and practical logistical supports during their diagnostic journey and in ongoing care. | Establish a Tasmanian Rare Disease Support Group. | Existing support groups on mainland Australia could establish a presence in Tasmania, or establish expertise and services specifically for rural people. |

**Figure 2.**
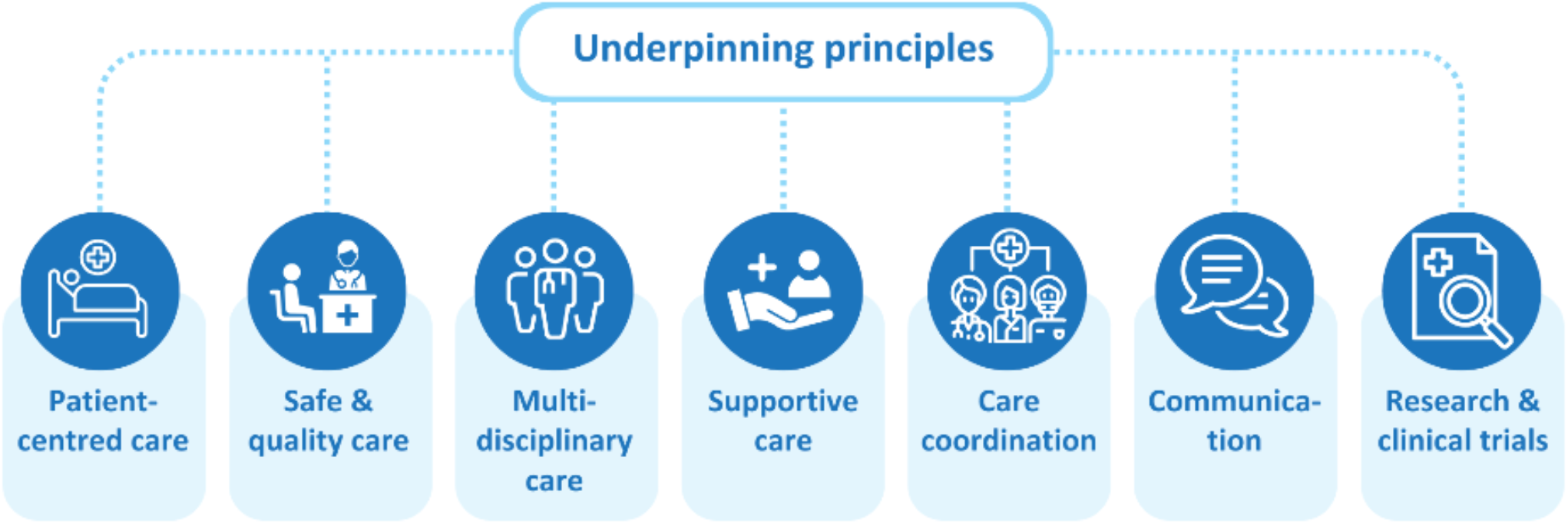
Underpinning principles relevant to a Rare Care Centre. Adapted from Cancer Australia^31^

The Centre would meet established needs for clear, Statewide referral pathways, so that a rural family physician who suspects mitochondrial disease, for example, knows what to do without hunting for protocols – the single port-of-call being the Rare Care Centre. The Centre model aims to reduce long waiting lists and duplicative tests through multidisciplinary liaison. From the consumer perspective, a single ‘hub’ with dedicated consumer liaison could help rare disease families connect with national registries, clinical trials, academic studies, and virtual support networks.

Healthcare consumers using services typically instituted in rare care centres report high satisfaction, improved treatment compliance, fewer clinic visits, and lower health system costs.^21-23^ The successful Rare Care Centre in Perth, Western Australia, provides holistic care to families affected by rare disease.^24^ While rare diseases are typically clinically distinct, with vastly different medical needs, they share many common characteristics including a prolonged diagnostic odyssey,^8^ concomitant care from multiple diverse clinicians,^25^ and challenges with healthcare communication.^26^ Families affected by rare disease live with uncertainty, isolation, caregiver burden, and mental health concerns. It is disruptive to all aspects of daily life. A Rare Care Centre, with its ability to provide wrap-around psychosocial, mental health, social, and school supports as well as improved access to clinical care, could significantly change the health dynamic for families with rare disease.^24^

It should be acknowledged that even with such a Centre, there would be a residual unmet need among healthcare consumers for whom a clinical geneticist or other clinician with an interest in genetics thinks a genetic or genomic test is clinically indicated, and the consumer wants the test, but the test is not accessible. Furthermore, future primary and implementation research should focus on improving the education and training of the non-genomic health professional workforce in relation to rare and genetic diseases, increasing the equity of access to genetic and genomic testing, finding ways to direct genomic outreach services to less served populations, and how to address funding challenges within genetic and genomic testing. A Centre would sit within the Tasmanian Health Service, the state’s public hospital and associated services system, and be designed to help Tasmanians find reliable information, navigate referral pathways and build clinician knowledge. An ideal structure would provide user-friendly information portals (online and via telehealth) where families and family physicians can quickly find credible rare disease resources. Integral to this structure, particularly in the Australian setting, would be access for clinicians from remote areas to targeted education and mentorship programs linked to the Centre, allowing non-genetics practitioners to gain confidence in genomics as part of routine diagnostic care.

High quality national data on congenital anomalies, including those meeting clinical criteria for rare disease, is currently unavailable. A state-based register for congenital anomalies was established in Western Australia in 1980 using active and passive monitoring,^27^ and may represent a model that Tasmania could implement. This could form a part of the Rare Care Centre with an aim to reduce the time to diagnosis and provide data for service improvement. Part of such a register could include addressing hospital coding limitations^28^ and the potential value of ORPHAcoding.^29^

The unexpectedly high response to our mixed methods research project warrants some discussion as it highlights the importance of rare disease care to the Tasmanian population. While individually rare, conditions that comprise rare and genetic diseases are collectively common: with a combined prevalence of 8%, they are more common than diabetes mellitus (6% prevalence in Tasmania).^30^ The results from this project place emphasis on the importance of awareness of the rare disease community. The establishment of a Rare Care Centre in Tasmania has the potential to significantly improve outcomes for people with rare and undiagnosed diseases.

Limitations of this study include the aggregated data analysis, which was chosen to preserve anonymity of the participants. A large proportion of the participants were healthcare consumers and their families and caregivers, which has provided rich data on their perspective, however participation in the healthcare professionals survey was limited and we therefore cannot draw strong conclusions on the views of non-genomic health professionals. We are planning follow-up studies to capture the voice of these professionals to help shape Tasmania’s Rare Care Centre.

## CONCLUSION

This paper presents stakeholder perspectives of rare disease diagnostic pathways in Tasmania, a population residing in regional, rural, and remote Australia. The evidence gathered by the researchers indicates that challenges and barriers to rare disease care are typically, but not exclusively, related to remoteness, and highlight the need for greater education and awareness of rare disease healthcare throughout consumers and practitioners in Tasmania. The primary recommendation forthcoming from this study is the establishment of a Tasmanian Rare Care Centre, to service the rural community. Unexpectedly high participation in this study highlights a call for help from the rare disease community, to be heard, and to be collaboratively cared for. With results that are conveyable to other rural settings, the rare disease community has spoken. It’s up to us to listen.

## Data Availability

Aggregated data from this study are available upon reasonable request to the authors.

## Acknowledgement statement

This research was funded by an Implementation Projects Grant from Australian Genomics’ National Health and Medical Research Council Grant GNT2000001.

## REFERENCES

1. Baynam G, Hartman AL, Letinturier MCV, et al. Global health for rare diseases through primary care. Lancet Glob Health. 2024; 12(7): e1192–e1199.

2. The Lancet Global Health. The landscape for rare diseases in 2024. Lancet Glob Health. 2024; 12(3): e341.

3. Wang CM, Whiting AH, Rath A, et al. Operational description of rare diseases: a reference to improve the recognition and visibility of rare diseases. Orphanet J Rare Dis. 2024; 19(1): 334.

4. Australian Government Department of Health and Aged Care. What we’re doing about rare diseases. 2022. 7-Nov-2022. https://www.health.gov.au/topics/chronic-conditions/what-were-doing-about-chronic-conditions/what-were-doing-about-rare-diseases

5. Cohen ASA, Berrios CD, Zion TN, et al. Genomic Answers for Kids: Toward more equitable access to genomic testing for rare diseases in rural populations. Am J Hum Genet. 2024; 111(5): 825–832.

6. Involve Australia. Guidelines for Community Involvement in Genomic Research. 2023:

7. Casauria S, Collins F, White SM, et al. Assessing the unmet needs of genomic testing in Australia: a geospatial exploration. Eur J Hum Genet. 2025; 33(4): 496–503.

8. Faye F, Crocione C, Anido de Pena R, et al. Time to diagnosis and determinants of diagnostic delays of people living with a rare disease: results of a Rare Barometer retrospective patient survey. Eur J Hum Genet. 2024; 32(9): 1116–1126.

9. Involve Australia. Guiding Questions for Researchers. 2023:

10. Sisk BA, Kerr A, King KA. Factors affecting pathways to care for children and adolescents with complex vascular malformations: parental perspectives. Orphanet J Rare Dis. 2022; 17(1): 271.

11. Lunke S, Eggers S, Wilson M, et al. Feasibility of Ultra-Rapid Exome Sequencing in Critically Ill Infants and Children With Suspected Monogenic Conditions in the Australian Public Health Care System. JAMA. 2020; 323(24): 2503–2511.

12. Lunke S, Bouffler SE, Patel CV, et al. Integrated multi-omics for rapid rare disease diagnosis on a national scale. Nat Med. 2023; 29(7): 1681–1691.

13. Ball M, Bouffler SE, Barnett CB, et al. Critically unwell infants and children with mitochondrial disorders diagnosed by ultrarapid genomic sequencing. Genet Med. 2025; 27(1): 101293.

14. Mallawaarachchi AC, Fowles L, Wardrop L, et al. Genomic Testing in Patients with Kidney Failure of an Unknown Cause: A National Australian Study. Clin J Am Soc Nephrol. 2024; 19(7): 887–897.

15. Long S, Schofield D, Kraindler J, et al. The PreGen Research Program: Implementing Prenatal Genomic Testing in Australia-A Commentary. Aust N Z J Obstet Gynaecol. 2025:

16. Best S, Vidic N, An K, Collins F, White SM. A systematic review of geographical inequities for accessing clinical genomic and genetic services for non-cancer related rare disease. Eur J Hum Genet. 2022; 30(6): 645–652.

17. Braun V, Clarke V. Using thematic analysis in psychology. Qualitative Research in Psychology. 2006; 3(2): 77–101.

18. Braun V, Clarke V. Reflecting on reflexive thematic analysis. Qualitative Research in Sport, Exercise and Health. 2019; 11(4): 589–597.

19. Australian Bureau of Statistics. 2021 Census Community Profiles. 2021. Accessed 2-Nov-2024. https://www.abs.gov.au/census/find-census-data/community-profiles/2021/6

20. Australian Government Department of Health and Aged Care. National Strategic Action Plan for Rare Diseases. 2020. 26 February 2020. https://www.health.gov.au/resources/publications/national-strategic-action-plan-for-rare-diseases

21. A O. The Agrenska centre: a socioeconomic case study of rare diseases. PharmacoEconomics (Auckland). 2002:

22. Chou C, Wiredu SO, Imhof LV, et al. Health Services Interventions to Improve the Quality of Care in Rare Disease: A Scoping Review. medRxiv. 2024:

23. Van Groenendael S, Giacovazzi L, Davison F, et al. High quality, patient centred and coordinated care for Alstrom syndrome: a model of care for an ultra-rare disease. Orphanet Journal of Rare Diseases. 2015; 10(1):

24. Baynam G, Siffleet J, Abbott T, et al. Rare care - Cross-sector care coordination. Eur J Med Genet. 2025; 74: 104995.

25. Teutsch S, Zurynski Y, Eslick GD, et al. Australian children living with rare diseases: health service use and barriers to accessing care. World J Pediatr. 2023; 19(7): 701–709.

26. Molster C, Urwin D, Di Pietro L, et al. Survey of healthcare experiences of Australian adults living with rare diseases. Orphanet J Rare Dis. 2016; 11: 30.

27. Nembhard WN, Bower C. Evaluation of the Western Australian Register of Developmental Anomalies: Thirty-five years of surveillance. Birth Defects Res A Clin Mol Teratol. 2016; 106(11): 894–904.

28. Scanlon P, Ridler G, Say G, et al. Measuring the impact of rare diseases in Tasmania, Australia. Orphanet J Rare Dis. 2024; 19(1): 399.

29. Mazzucato M, Pozza LVD, Facchin P, et al. ORPHAcodes use for the coding of rare diseases: comparison of the accuracy and cross country comparability. Orphanet J Rare Dis. 2023; 18(1): 267.

30. Australian Bureau of Statistics. National Health Survey. 2022. https://www.abs.gov.au/statistics/health/health-conditions-and-risks/national-health-survey/2022

31. Cancer Australia. National Optimal Care Pathways Framework. 2024. https://www.canceraustralia.gov.au/sites/default/files/2025-01/national-optimal-care-pathways-framework.pdf

